# A cross-sectional study to evaluate access to antenatal care services in Twifo Hemang Lower Denkyira district of Ghana

**DOI:** 10.1101/2024.05.09.24307150

**Authors:** John Hammond, Silas Adjei-Gyamfi, Doreen Brew Daniels, Godfred Kwabena Sarpong, Hirotsugu Aiga, Tsunenori Aoki

**Affiliations:** School of Tropical Medicine and Global Health, Nagasaki University, Nagasaki, Japan; Central Regional Health Directorate, Ghana Health Service, Cape Coast, Central Region, Ghana; Savelugu Municipal Hospital, Ghana Health Service, Savelugu, Northern Region, Ghana; Elmina Polyclinic, Ghana Health Service, Elmina, Central Region, Ghana

**Keywords:** ANC attendance, ANC initiation, Antenatal care, Community-based Health Planning and Services, Pregnant women, Southern Ghana

## Abstract

**Background:** Antenatal care (ANC) which is an essential component of the reproductive, maternal, newborn, and child health continuum of care is found to positively correlate with supervised delivery and the reduction of maternal deaths. In Ghana, few studies have explored how ANC is influenced by the community-based health planning and services (CHPS) policy, and in the Central Region, evidence is non-existent. This study aimed to determine factors that influence access to ANC services provided through the CHPS policy in the Twifo Hemang Lower Denkyira district in the Central Region of Ghana.

**Methods:** A cross-sectional study examined 310 women aged 15-49 years, having children less than 12 months, and interviewed using a structured questionnaire. Univariate and multivariate logistic regression analyses were conducted using STATA 17 and results were reported as odds ratios at a confidence level of 95%.

**Results:** ANC coverage and proportion of early ANC initiation were 93.9% and 69.1% respectively.

Being unmarried (AOR=0.125, 95%CI=0.012,0.926), and home delivery (AOR=0.013; 95%CI: 0.001,0.176) were associated with decreased odds of at least one ANC visit during pregnancy. Larger (≥11) household size (AOR=3.848; 95%CI=1.914,16.21), lesser (<4) ANC contacts (AOR=6.332; 95%CI=2.049,19.57), and home visitation by CHPS staff (AOR=1.813; 95%CI=1.014,3.243) were associated with higher odds of late ANC initiation while average monthly income (AOR=0.123; 95%CI=0.024,0.630) was associated with reduced odds of late ANC initiation. Interestingly, knowledge about ANC and pregnancy, and geographical variables like receiving ANC services from CHPS zones, and distance to CHPS zones were not statistically significant with either ANC attendance or time of ANC initiation after controlling for the effect of other variables.

**Conclusion:** Though ANC and early ANC initiation coverages were relatively high, the complexities in the given correlates of ANC accessibility require a multi-sectoral approach to strengthen community-based services to increase the survival of pregnant women and unborn babies.

## Introduction

The International human rights law recognizes the right of women and adolescent girls to survive pregnancy and childbirth through the provision of appropriate sexual and reproductive health services (1). According to the World Health Organization (WHO), access to adequate healthcare must be provided with sufficient and appropriate content (2). However, globally, over 400 million people lack access to primary care (3), including pregnant women.

Antenatal care (ANC), which is the care provided by skilled healthcare providers to women during pregnancy for positive birth outcomes, forms an essential part of the reproductive, maternal, newborn, and child health (RMNCH) continuum of care. ANC has become a global priority over the past decade intending to increase its coverage (4); as such, between 2015 and 2020, about 87% of pregnant women had at least one ANC visit with a skilled care provider globally (5). Also, in sub-Sahara Africa (SSA), the proportion of women who had at least one ANC visit during pregnancy has improved from 69% in 2006 (6) to 78.5% in 2021 (7). RMNCH services like ANC does not only reduce maternal and child mortalities (8,9) but also prevent adverse birth outcomes like low birthweight and preterm births (10,11), especially in poorly resourced countries (12). Early identification of high-risk pregnancies through ANC leading to complications like severe bleeding, infections, pre-eclampsia/eclampsia, unsafe abortions, which contribute to nearly 80% of all maternal deaths, and the provision of appropriate care by skilled health personnel at all levels of the continuum of care has been proven to be the best preventive system to save lives of women and children (9,13–15).

The challenges women face in accessing skilled care while pregnant through to delivery in poor areas have been reported as a significant contributor to maternal deaths (16). While 17% of global births occurred in SSA, it contributed to the highest proportion (57%) of the 358,000 deaths that occurred due to pregnancy and childbirth (17). Most studies have attributed ANC accessibility to factors including socio-culture, socio-economic, and socio-demographic (religion and rural/urban residence) factors among others, as predictors of timely and continuous ANC attendance (8,18,19). It, therefore, underscores the need for comprehensive and sustainable healthcare policies to address these gaps, creating inequities in access for women, especially those in rural communities (20).

Over the past two decades, Ghana has been implementing the community-based health planning and services (CHPS) policy, aiming to reduce access barriers to healthcare through the provision of primary health care (PHC) services at the community level. The primary strategy of the policy is the posting of a community health nurse/officer (CHN/O) in a clearly defined area with the support of the community leadership and support system to plan and mobilize resources to provide services (21,22). The initiative, which is the primary strategy for the provision of community-focused services, has contributed to a significant drop in maternal and child deaths and the increasing performance of ANC and vaccination services, particularly in rural areas (23). Despite its promising benefits in Ghana, 2.3% of women did not attend ANC before delivery (24), 22% of all deliveries were assisted by unskilled personnel (25), and only 9.9% of births occurred within 8 km of CHPS zones (26), and this could partly explain why Ghana did not meet the Millenium Development Goal’s target on maternal and infant health.

In the Central Region of Ghana, CHPS facilities which serve 52% of rural populations in the region, contribute to about 16% of all ANC registrants and 8% of skilled deliveries (27). The Twifo Hemang Lower Denkyira (THLD) district is one of the rural districts with poor maternal health indicators despite the increased investment in scaling up PHC services closer to the doorsteps of many more households and communities. In 2020, ANC coverage and first-trimester ANC registrants were 56.4% and 57.7% respectively, with only 32.5% of pregnant women having access to skilled delivery care (27), indicating a gap between ANC and skilled delivery, as seen in most SSA countries. There is a need to understand how pregnant women in rural settings interact with the healthcare system and what influences their access to ANC in the context of CHPS. Considering several pieces of evidence that make CHPS a very promising policy for the improvement of services, particularly for women, access to healthcare in the district remains poor. Only a few studies have examined the access to ANC services provided through the CHPS strategy for women in the region and its impact on achieving quality maternal and child healthcare. This study therefore aimed to explore factors that influence women’s access to ANC services provided through the CHPS strategy in the THLD district of the Central Region of Ghana.

## Materials and Methods

### Study setting

The study was conducted at THLD district which is one of the districts among the 22 districts in the Central region of Ghana, with its administrative capital at Twifo Hemang. It stretches for about 75km from the regional capital, Cape Coast, and lies between latitudes 5° 50’ N and longitude 1° 50’ W, located in the north-western part of the region with a total landmass of 674 sq. km (Figure 1). The estimated population in 2020 was 67,341, of which 51% are females, with an estimated 4% (2,694) expected to be pregnant and a population density of about 82 per square kilometer (28). Whereas the number of women of reproductive age in 2020 was 16,073, the total ANC and postnatal visits were 7,502 and 1,324 respectively. THLD district is predominantly rural (86%) with 300 settlements and 84 clearly defined communities. Most of the remaining settlements are farmsteads, usually with a population below 500 people. The health delivery system in the district is made up of the orthodox and traditional systems, with the latter consisting of traditional birth attendants, herbalists, fetish priests, and spiritualists, and is thought to play a crucial role in the health-seeking behavior of most people in the district. Health delivery under the orthodox system in the district exists up to two levels (A and B) under the country’s PHC system, that is, the community and sub-district levels. It is divided into four sub-districts with no district hospital but has three health centres, one community clinic, and 13 functional CHPS zones (27) as shown in Figure 1.

**Fig 1.**
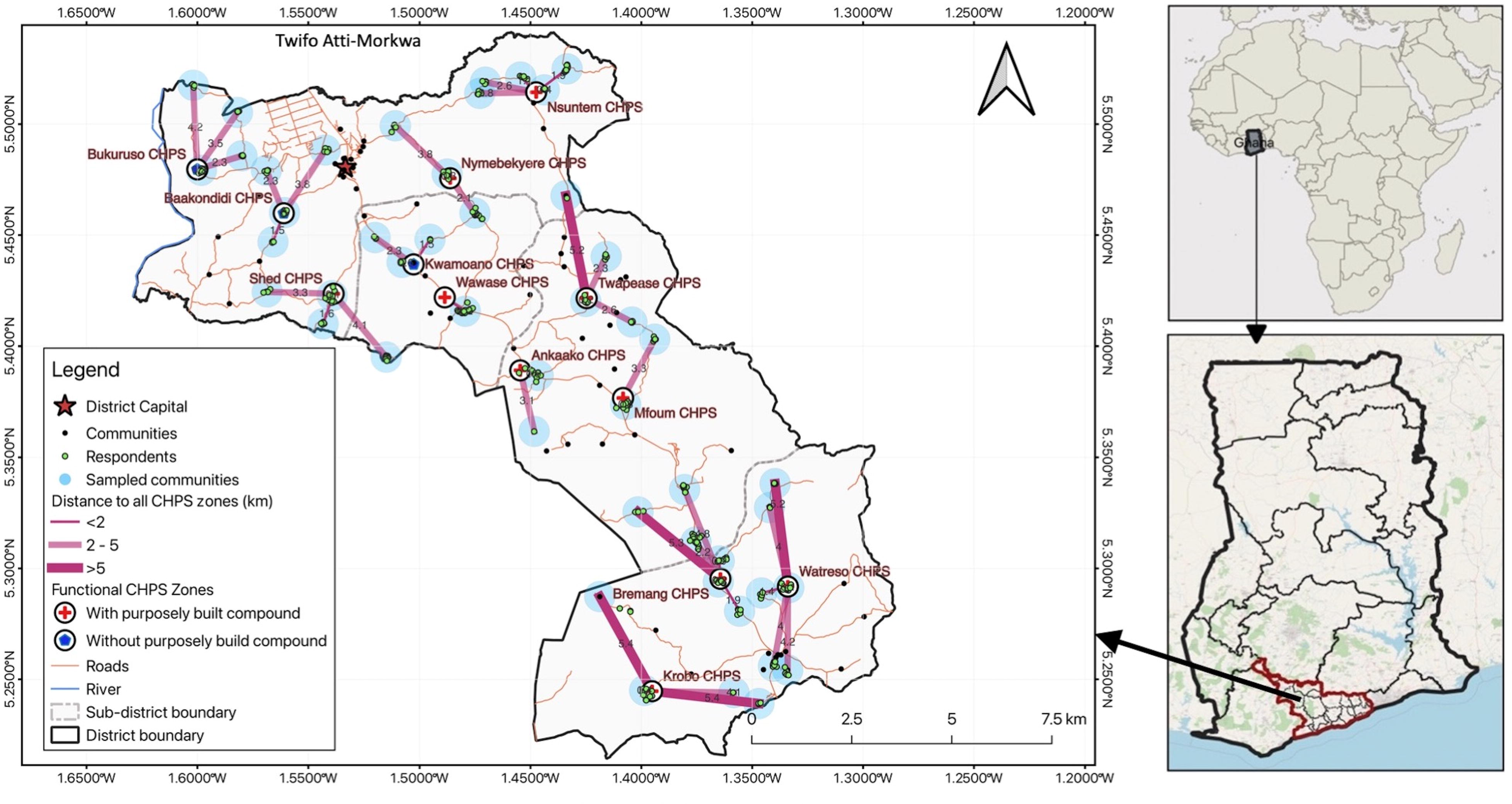
Map of the THLD district showing communities and measured distance to all CHPS zones.

### Study design and population

To assess access to ANC services in the THLD district of the Central Region under CHPS policy, a descriptive cross-sectional quantitative study was used. This study targeted women of reproductive age (15-49 years) having at least one child aged 12 months or younger preceding data collection, living or residing within communities of all the demarcated CHPS zones of the district.

### Sample size

The sample size (N_0_) was determined using Cochran’s (1977) formula; N_0_ = [{(Zα/2)^2^(1–p)p}/d^2^]. Where “N_0_” is the estimated sample size, “Zα” is the confidence level at 95% (standard value of 1.96), “p” is the estimated proportion of women accessing maternal healthcare services in CHPS zones in Ghana (76%) (18), and “d” is the level of precision/margin of error of 0.05. Hence, N_0_ = [{(1.96)^2^ x (1 – 0.76) x 0.76}/(0.05)^2^] = 280.3 ∼280. With a minimum sample of 280, a non-response rate of 10% was added to attain an estimated final sample size of 310.

### Recruitment method

During sampling, all 13 CHPS zones in the THLD district were selected for the study. Probability proportional to size technique was used to determine the sample sizes for each CHPS zone. Using the vaccination records in the child welfare clinic register in each CHPS zone as the sampling frame (29), a total of 1751 eligible participants (children born under 12 months) were obtained. Thereafter, a simple random sampling method was used to select the required sample size of 310 from the CHPS zones and the selected children were directly linked to their mothers and sorted according to the community of residence. In total, 48 communities were obtained and the addresses (including telephone numbers) of the mothers were taken for identification at the community level. Within the community, research assistants were supported by the CHN/Os and the community volunteers to trace the mother-child dyads. The respondents were interviewed in their place of residence after booking an appointment with them and signing an informed consent/assent form. In the absence of a study respondent at the time of the visit, the research assistant rescheduled the interview to a later time.

### Data collection

The data collection was conducted by five trained research assistants from 15^th^ February to 15^th^ March 2022. Data was collected using a pretested standardized questionnaire with the aid of mobile data collection software (Open Data Kit Collect-version 2021.2.4). The information on socio-demographic, socioeconomic (household assets), antenatal, and CHPS location/distance characteristics as well as respondents’ knowledge of ANC and pregnancy were collected through face-to-face interviews with participants using the local dialect (Fantse and Twi). Other relevant information on ANC services was confirmed from maternal and child health record books. Before each session of the interview, global positioning system (GPS) coordinates were obtained from respondents which were used to estimate distances from the respondent’s place of residence to the nearest CHPS zone.

### Measurement of study variables

A description of the key variables that were collected by the research assistants is shown in Table 1. The outcome variables in the study were At least one ANC attendance and late ANC initiation. Whereas the exposure variables included all maternal background (sociodemographic, socioeconomic, and antenatal variables) and CHPS location/distance characteristics.

### Data analysis

Data analysis was conducted using STATA version 17 (Stata Corporation, Texas, USA) at a significance of p<0.05. Simple descriptive statistics were performed on categorical variables and summarized into frequency distributions and percentages. Means and standard deviations were calculated for continuous variables and were further categorized based on the existing literature. ANC attendance was coded as “0” for “No ANC attendance” and “1” for “ At least one ANC attendance”; while the timing of ANC initiation was also coded as “0” for “Early ANC initiation” and “1” for “Late ANC initiation”. Socioeconomic characteristics were measured with principal component analysis to construct household wealth terciles (poor, middle, and rich) using household assets owned by respondents (30). Knowledge was assessed using composite scores of responses about ANC and pregnancy, with each correct answer given “1” mark and “0” for a wrong answer. A median score was generated from the total scores, and participants who scored equal to or above the median score were classified as having adequate knowledge, and those below the median score were classified as having inadequate knowledge (10,29,31). All GPS coordinates collected from participants were processed using QGIS software for analysis through the Euclidean technique to establish distance from the respondent’s place of residence to the CHPS zones (32,33).

Bivariate analyses (Chi-square/Fisher’s exact test) were used to explore the association between outcome variables and all background (exposure) variables. Simple logistic regression analyses were further conducted to confirm the bivariate associations. Based on previous studies, significant predictor variables in the bivariate association that are plausible and relevant in explaining the outcome variables were forwarded into the multivariate logistic regression model after controlling for multicollinearity. During multicollinearity testing via variance inflation factor (VIF), the variables with a VIF less than 5 were chosen for the final logistic analyses (11). Multivariate logistic regression was used to identify factors for at least one ANC attendance or late ANC initiation and the analysis results were reported as odds ratios with its corresponding 95% confidence interval.

### Ethical declarations

Ethical approval was sought from the Nagasaki University Ethics Review Committee and Ghana Health Service Ethics Review Committee with protocol approval numbers NU_TMGH_2021_195_1 and GHS-ERC: 023/01/22, respectively. Additionally, written permission was obtained from the Central Regional and THLD District Health Directorates. Opinion leaders in the selected communities were informed about the presence of the research team. Only participants who agreed to participate in the study were made to sign the written informed consent after explaining the content to them in the presence of their witnesses. Also, written informed assent was signed by parents/guardians/legal representatives of participants (minors) who are less than 18 years old.

## Results

### Sociodemographic characteristics of respondents

Of the 310 recruited mother-child dyads who fully responded to the study, 24.2% were between the ages of 30-34 years, with a mean(±sd) age of 27.6(±6.9) years. Most of the respondents were married (78.1%), were engaged in some form of occupation (80.6%), belonged to the Christian religion (95.5%) and Fantse ethnic group (47.4%), and had a parity of 2 to 4 births (46.1%). A greater proportion of these respondents (87.1%) and their partners (86.1%) attained some form of formal education with secondary/higher as their highest educational level. More than half of the respondents had a household size between 6-10 people (52.9%) while slightly above one-third (35.5%) of them had their youngest child between the ages of 9 to 12 months (Table 2).

### Socioeconomic and antenatal characteristics of respondents

Most of the respondents were enrolled in the national health insurance scheme (91.6%) and had a higher knowledge level about ANC and pregnancy (96.5%) while 42.3% of them were classified as poor based on their household wealth index. Despite the free maternal healthcare policy in Ghana, all the women claimed they paid some money for receiving ANC services from the health facilities with 26.5% of them paying between Ghc 1.0–5.0 (USD 0.1–0.5).

ANC coverage was nearly universal (93.9%) with most of them accessing ANC service from the CHPS zones (71.8%). In addition, 69.1% of the women initiated ANC services at an early stage of pregnancy (thus within the first trimester) while 12.6% of them made less than four visits before delivery. The decision to attend ANC was mainly made by most women themselves (77.4%) and almost one-third of the women were escorted by their partners (32.0%). Although 34.8% of the respondents received home visits from CHPS zone staff, the prevalence of home delivery was 19.7% (Table 3).

### Distance and location of CHPS zones

Table 3 describes the distance and location of CHPS zones (health facilities) for ANC services. More than half (52.6%) of the respondents resided in communities without health facilities, and 80.7% reported that the CHPS compound is the nearest health facility to them. However, 15.5% of the nearest CHPS zones were uncompleted functional zones. About 81.6% of the women travel for less than 30 minutes before getting to the nearest CHPS zones, with walking (46.5%) and use of commercial motorbikes (34.8%) as the primary mode of transport. Also, 90.3% of the women spent between GHc 1.0–10.0 (USD 0.1–1.1) to and from the nearest health facility per ANC visit on roads described by most as feeder/dusty (56.1%). The distance measured from the place of the residence revealed that about 80.7% of the women traveled less than five kilometers to the nearest functional CHPS zone (Figure 1).

### Bivariate association of antenatal accessibility with background variables

As displayed in Tables 1, 2, and 3, marital status (p=0.028), mother’s educational level (p=0.003), partner’s educational level (p=0.043), mother’s health insurance status (p=0.040), household wealth index (p=0.003), knowledge about ANC and pregnancy (p<0.001), place of delivery (p<0.001), receiving home visit from CHPS zone staff (p=0.007), health facility in the community (p<0.001), CHPS zone functionality status (p=0.008), and distance to nearest functional CHPS zones (p=0.046) exhibited bivariate correlation with at least one ANC attendance. On one hand, household (family) size (p=0.030), mother’s average monthly income (p=0.017), household wealth index (p=0.029), frequency of ANC visits (p<0.001), receiving home visit(s) from CHPS zone staff (p=0.049), CHPS zone functionality status (p=0.002), distance to nearest functional CHPS zones (p=0.012) were significantly associated with late ANC initiation at the bivariate level.

### Multivariate analysis on associated factors for antenatal accessibility

After registering no multicollinearity issues among the eleven and seven predictor variables that showed significant bivariate association with at least one ANC attendance and late ANC initiation respectively, two (marital status, place of delivery) and four (household size, mother’s average monthly income, frequency of ANC visits, receiving home visit from CHPS zone staff) predictor variables respectively remained statistically significant at the multivariate analysis level (Table 4).

The findings revealed that unmarried women were 98.7% (=[1–0.013]x100) less likely to attend ANC services compared to married ones (AOR=0.013; 95%CI=0.001,0.176; p=0.001). Women who delivered at home had lower odds of attending ANC during pregnancy than those who delivered in a health facility (AOR=0.125; 95%CI=0.012,0.926; p=0.047).

On the other hand, women with a larger household size of more than 11 people were 3.848 times more likely to initiate late ANC services than their counterparts (AOR=3.848; 95%CI=1.914,16.21; p=0.046). The likelihood of a pregnant woman who made less than four ANC visits to initiate late ANC services is 6.332 times higher than those who made at least four ANC visits during pregnancy (AOR=6.332; 95%CI=2.049,19.57; p=0.001). Additionally, pregnant women who received home visits by CHPS staff had 1.813 times increased odds of late ANC initiation compared to those who did not receive home visit sessions (AOR=1.813; 95%CI=1.014,3.243; p=0.045). Finally, compared to women who had no income, those whose average monthly income ranged between GHc 500–1000 (USD 56–111) were 87.7% (=[1–0.123]x100) less likely to initiate ANC services in the second or third trimester of pregnancy (AOR 0.123, 95%CI=0.024,0.630; p=0.012).

## Discussion

The study examined factors that influence access to ANC services in rural communities within the THLD district of the Central Region of Ghana in the context of CHPS policy implementation. Findings from the study showed that 93.9% of the respondents had at least one ANC service from a skilled provider, with 69.1% initiating their first visit within the first trimester of pregnancy. This is higher than the regional ANC coverage and proportion of first-trimester visits of 81% and 53% respectively (27) but agrees with the general high coverage in Ghana as reported by several studies (24,34). In contrast, other studies in Ghana (35) and Rwanda (36) reported only 43.1% and 25% of early ANC initiation respectively. In our study, 93% of the women also made four or more ANC visits before delivery which is higher than the Ghana national average of 89% (24) and rates in other developing countries (29,37,38). This is an indication of how the interest of Ghanaian women has developed in the benefits of attending ANC, which could be attributed to the heightened efforts to expand health facility coverage to improve access to healthcare, especially in bringing quality healthcare services close to the people (39).

Marital status offered significant protection for ANC attendance, as unmarried women had reduced odds of making at least one ANC visit. This finding is consistent with many other studies conducted in Ghana (8,31,40,41), and Kenya (38), but contradicts the study results from Ethiopia (42,43). The possible reason for this finding could be attributed to society’s values on marriage and pregnancy. For instance, in most rural Ghanaian communities, unmarried women are not to engage in sexual acts (8). In a situation when pregnancy occurs, most of these women tend to hide their pregnancies until it is obvious to avoid being stigmatized and ostracized by society, which could contribute to reducing their ANC contacts or failing to attend ANC at all (31,38,44,45). On the other hand, married women are seen to have a higher possibility of receiving support from their partners (husbands) during pregnancy (8) which increases their chances of higher ANC visits.

The place of delivery had a significant influence on ANC attendance, as women who delivered at home were less likely to make at least one ANC visit. This study’s results agree with similar studies conducted in most developing countries (35,46,47). In Ghana, there is increased awareness creation (via health education, and counseling) on services for pregnant women, free maternal healthcare under the NHIS, as well as the expansion of CHPS, which is helping to bridge the access gap for vulnerable populations (8,48). Notwithstanding, the perceived mistreatment of pregnant women by healthcare providers (49,50) could be responsible for the home deliveries and low ANC visits. It is also worth noting that, despite the availability of at least one midwife to offer skilled care for pregnant women in almost all the CHPS zones in the district, most women are demotivated to visit ANC for service due to some cultural and personal beliefs (27).

Household size of more than 11 people was a significant correlate of late ANC initiation. In congruence with similar studies across the globe (51–54), having a larger household size is a strong predictor of late ANC initiation. Our study results contradict findings in Ethiopia (55) and Northern Ghana (45) which concluded that women with 4–9 living children were 3.5 times more likely to initiate ANC early. Household sizes in rural communities are mostly larger than those in urban communities (24), as more than half of our study respondents (57.1%) had a household size of 6 or more people. In the Ghanaian context, there are numerous societal responsibilities and pressures conferred on women with larger household sizes, as these women serve as the gatekeepers and managers of rural homes (8). This can affect the time and resources the pregnant woman may require to start and sustain ANC and, in most cases, prioritize their farming or trading activities to provide for their families. Also, in some instances, those women who have experienced successful pregnancies may develop some level of confidence that they can survive the early period of pregnancy without any skilled assistance leading to late initiation (45,54,56).

Women who made less than four ANC visits were more likely to initiate their first ANC after the first trimester of pregnancy which is consistent with some study findings (57,58). Evidence has shown that women who initiate early ANC have higher odds of receiving four or eight ANC contacts than those with late ANC initiation (7,45,59). Higher ANC contacts and/or early ANC initiation is essential as it ensures that pregnant women get all the required content and higher quality ANC services at the right time (14,58). Most women in rural communities have unplanned pregnancies and may not be aware of being pregnant until they feel unwell before visiting the health facility, which is usually late. Furthermore, other sociocultural and religious beliefs and misconceptions surrounding pregnancy within the Ghanaian communities potentially discourage the use of orthodox healthcare services (60,61) which may lead to late or no ANC visits. Reportedly, low economic status, perception of no problem with pregnancy, and misinterpretation of recommendations from healthcare providers (45,62,63) could ignite late ANC initiation leading to non-achievement of the recommended number of visits before delivery.

Home visit by health staff was independently associated with the time of ANC initiation. Home visits form a significant part of the daily activities of health staff in CHPS zones. This house-to-house visitation enables individuals and households to receive appropriate information and care at their doorstep throughout the RMNCH continuum of care (22,64). Our study revealed that pregnant women were found to have higher odds of attending ANC late if they received a home visitation from health staff from the CHPS zone. This finding is comparable with a randomized control trial in Nigeria (65) but contradicts the results of a similar study by Kumbeni and his colleagues in the Upper East region of Ghana (40) which reported that women who received at least one home visit had higher odds of initiating early ANC and achieving at least eight ANC visits. The plausible reason for this finding may be due to the trust built between the pregnant woman and the healthcare professionals through home visit sessions (40,48). These women misunderstand the services they received during the home visitation as the same as the facility care. Consequently, they become self-satisfied and do not find it appropriate to go to the health facility, especially when they feel nothing is wrong with them (45).

Monthly income offered significant protection for ANC initiation, as women with an average income between GHc 500–1000 had reduced odds of initiating late ANC services. This finding agrees with most studies in the SSA (51,52,66), which established a substantive relationship between women’s income level and initiation of ANC. High monthly wage offers these women enormous leverage to overcome late visitation (67,68). Most Ghanaian women are unable to afford other indirect costs of accessing ANC (transportation, laboratory investigations, and other medications), despite the free maternal healthcare under the NHIS, making women who earn more income better placed to afford these services (66,69), and avail themselves for early ANC services. Nearly 77% of the study’s respondents were found to be self-employed, with most (39.4%) being farmers. Thus, their income is not regular and often not even close to the Ghanaian minimum wage due to the unstable economic situation in Ghana. The apparent financial constraint forces these women to adopt options to reduce expenditure, including delaying ANC initiation (70). This gives credence to the importance of pro-poor policies that can augment the financial situation of these women and ultimately assist them in making appropriate reproductive health decisions.

### Strengths and limitations of the study

This is one of the few studies in Ghana that examined access to ANC by pregnant women in rural communities in the context of CHPS; thus, the findings provide valuable insights for policy strengthening. Moreover, the methodology used allowed for the gathering of primary data, which characterizes the uniqueness of the study setting. Restricting participants to only 12 months post-partum mothers preceding data collection reduced possible recall bias.

Nonetheless, the type of study design used may have limited the impact of the study due to the interpretation of the results as it could only allow for inferences but not causal relationships. Although there were efforts to limit information bias, the self-reporting used may have introduced recall and reporting biases. For instance, some mothers refused to disclose information on their monthly income, and others were not sure about their earnings. Additionally, since only mother-child dyads were used, information about mothers with adverse birth outcomes like perinatal deaths was not collected which might have affected study results. The use of the Euclidean distance to measure the distance of respondents to CHPS zones did not consider other topographical factors such as terrain. However, this method, though not sophisticated, has been an acceptable method for determining accessibility. Finally, only functional CHPS zones were used as the unit of measure, even though some of the respondents were closer to other types of health facilities which may have limited the data as further studies may be required to bridge that knowledge gap.

## Conclusion

ANC coverage and early ANC initiation rate in the district were found to be high. Being unmarried, and home delivery were associated with at least one ANC visit. Independent correlates of late ANC initiation included larger household (family) size, fewer ANC contacts, home visitation by CHPS staff, and maternal average monthly income. Although a greater proportion of respondents received ANC services from CHPS zones, resided closer to CHPS zones, and had adequate knowledge about ANC and pregnancy, these expected plausible factors were not statistically associated with ANC attendance and time of ANC initiation.

The interaction of these given factors requires a multi-sectoral approach to mitigate them. The Government of Ghana through its relevant agencies including Ghana Health Service (GHS), should intensify the implementation of pro-poor policies such as Livelihood Empowerment Against Poverty (LEAP) which provides financial support for the poor, free maternal healthcare services, fertilizer subsidy for farmers, free education with more emphasis on girl education, among others, so that the livelihood of these vulnerable women in rural communities could be improved.

Additionally, the Ghana Ministry of Health (MoH) and stakeholders, including the Ministry of Education, Local Government Ministry, and other Non-Governmental Organizations should ensure more women in rural communities take advantage of the free education policy, thereby empowering them to make informed decisions on their reproductive rights.

The provision of accurate, appropriate, and timely information can potentially alter health-seeking behaviors in the district. Thus, the MoH and GHS should strengthen Behavior Change Communication strategies provided through health promotion and education. All possible avenues to share this information should be explored, including the use of community health management committees, community health volunteers, schools, community support groups, and community opinion leaders.

The MoH and GHS should also strengthen policies on home visits, including resourcing health facilities appropriately to gain the needed positive results. The content should be re-examined with a more innovative approach to providing the service and appropriate follow-up where necessary. Further qualitative studies need to be conducted to provide more insights into how these factors affect access to ANC and its impact on the individuals, households, and communities in the THLD district.

## Data Availability

The datasets collected, generated, and/or analyzed during the present study are available from the corresponding author upon reasonable request.

## Acknowledgments

The authors are very thankful to The Project for Human Resource Development Scholarship - Japan International Cooperation Agency (JICA); Nagasaki University School of Tropical Medicine and Global Health – Japan; and Central Regional Health Directorate, Ghana Health Service – Ghana, for supporting this study. Our gratitude also goes to all data collectors, research respondents, and volunteers who greatly contributed to this study.

## Abbreviations

ANC: Antenatal Care
CHN/O: Community Health Nurse/Officer
CHPS: Community-based Health Planning and Services
GHc: Ghana cedis
GHS: Ghana Health Service
GPS: Global Positioning System
MoH: Ministry of Health
PHC: Primary Health Care
RMNCH: Reproductive, Maternal, Newborn, and Child Health
SSA: Sub-Saharan Africa
THLD: Twifo Hemang Lower Denkyira
USD: United States of America Dollars
VIF: Variance Inflation Factor
WHO: World Health Organization

## Consent For Publication

Not applicable

## Authors’ contributions

JH, SAG, DBD, and TA designed and conceptualized the study. JH, SAG, and DBD carried out the data collection as GKS, HA, and TA played supervisory roles. JH, SAG, GKS, HA, and TA performed the data analysis and interpretation. JH, SAG, and DBD drafted the initial manuscript. All authors reviewed and approved the final manuscript.

## Authors’ acronyms

John Hammond (JH); Silas Adjei-Gyamfi (SAG); Doreen Brew Daniels (DBD); Godfred Kwabena Sarpong (GKS); Hirotsugu Aiga (HA); Tsunenori Aoki (TA)

## Supporting information

S1 Figure 1. Map of the THLD district showing communities and measured distance to all CHPS zones

S1 Table 1. Description of key variables

S2 Table 2. Sociodemographic characteristics of respondents

S3 Table 3. Socioeconomic and antenatal characteristics of respondents

S4 Table 4. Distance and location of CHPS zone characteristics

S5 Table 5. Logistic regression on associated factors for antenatal accessibility

