## Supplementary material for "A cross-sectional study to evaluate access to antenatal care services in Twifo Hemang Lower Denkyira district of Ghana": Tables

**Table 1: Description of key variables**

| **Variable** | **Definition and description** | **Variable type** | **Categories used** |
| --- | --- | --- | --- |
| ANC attendance | The ability of respondents to make at least one ANC visit during pregnancy | Binary | At least one ANC visit; No ANC visit |
| ANC initiation | Timing of the first ANC visit before delivery of the youngest child ≤12 months and dichotomized into early initiation (0–12 weeks of gestation or first trimester) and late initiation (≥13 weeks of gestation or second and third trimester) | Binary | Early ANC initiation; Late ANC initiation |
| Age group | Age at last birthday in years and categorised | Ordinal | 15–19; 20–24; 25–29; 30–34; 35+ |
| Marital status | Having customary or legal partner | Binary | Married; Not married |
| Religion | Religious affiliation of the respondent | Binary | Christianity; Islam |
| Ethnicity | Belonging to any tribal or ethnic group | Nominal | Fantse; Denkyira; Ewe; Akuapim; Others |
| Educational level | The highest level of education attained by respondents and partners | Nominal | No formal education; Primary; Secondary/ higher |
| Household/family size | The number of people living within the household of respondents. | Ordinal | 1–5; 6–10; 11+ |
| Parity | The total number of children born by respondent | Ordinal | 1; 2–4; 5+ |
| Age of child | The age of the youngest child in completed months verified with the child vaccination records. | Ordinal | ≤4; 5–8; 9–12 |
| Employment | The status of occupation of the respondent and the partner | Binary | Unemployed; Employed |
| Household wealth index | Constructed using household items possessed by respondents and analyzed using principal component analysis before grouping them into terciles. | Nominal | Poor; Middle; Rich |
| Place of ANC attendance | Type of facility where respondent received ANC care during pregnancy. | Nominal | CHPS; Clinic; Health centre; Hospital/ Polyclinic |
| Frequency of ANC visit | Number of ANC visits before delivery and categorized into <4 and ≥4 visits. | Binary | <4; ≥4 |
| Decision to attend ANC | Who decides for the respondent to attend or not to attend ANC during her pregnancy. | Nominal | Myself; Husband/ partner; Others |
| Knowledge | Participants' level of knowledge about ANC and pregnancy and categorised into Inadequate and Adequate by assessing composited knowledge scores using a median cut-off point | Binary | Inadequate; Adequate |
| CHPS functionality status | The state of functionality of the CHPS zone according to the Ghana Health Service CHPS implementation guideline (2016). | Binary | Uncompleted; Completed |
| Distance to CHPS zone | The measured distance from participants’ place of residence to the nearest CHPS zones. | Binary | <5km; ≥5km |

**Table 2: Sociodemographic characteristics of respondents**

|  |  |  | **Bivariate analysis** | | | | | | |
| --- | --- | --- | --- | --- | --- | --- | --- | --- | --- |
| **Variables** | **Total (%)** |  | **ANC attendance (%)** | | **p-value^a^** |  | **ANC Initiation (%)** | | **p-value^a^** |
|  |  |  | **No**  **(n=19)** | **Yes**  **(n=291)** |  |  | **Early (n=201)** | **Late (n=90)** |  |
| **Age group** (mean:27.6; sd:6.9) | |  |  |  |  |  |  |  |  |
| 15-19 years | 47 (15.2) |  | 1 (5.3) | 46 (15.8) | 0.602 |  | 25 (12.4) | 21 (23.3) | 0.150 |
| 20-24 years | 65 (21.0) |  | 5 (26.3) | 60 (20.6) |  |  | 40 (19.9) | 20 (22.2) |  |
| 25-29 years | 69 (22.3) |  | 5 (26.3) | 64 (22.0) |  |  | 48 (23.9) | 16 (17.8) |  |
| 30-34 years | 75 (24.2) |  | 6 (31.6) | 69 (23.7) |  |  | 51 (25.4) | 18 (20) |  |
| 35+ years | 54 (17.4) |  | 2 (10.5) | 52 (17.9) |  |  | 37 (18.4) | 15 (16.7) |  |
| **Marital status** |  |  |  |  |  |  |  |  |  |
| Not married | 68 (21.9) |  | 8 (42.1) | 60 (20.6) | 0.028^*^ |  | 38 (18.9) | 22 (24.4) | 0.280 |
| Married | 242 (78.1) |  | 11 (57.9) | 231 (79.4) |  |  | 163 (81.1) | 68 (75.6) |  |
| **Religious affiliation** |  |  |  |  |  |  |  |  |  |
| Christianity | 296 (95.5) |  | 17 (89.5) | 279 (95.9) | 0.193 |  | 193 (96.0) | 86 (95.6) | 0.854 |
| Islam | 14 (4.5) |  | 2 (10.5) | 12 (4.1) |  |  | 8 (4.0) | 4 (4.4) |  |
| **Ethnicity** |  |  |  |  |  |  |  |  |  |
| Akuapim | 147 (47.4) |  | 2 (10.5) | 19 (6.5) | 0.753 |  | 95 (47.3) | 42 (46.7) | 0.623 |
| Denkyira | 73 (23.6) |  | 3 (15.8) | 70 (24.1) |  |  | 52 (25.9) | 18 (20.0) |  |
| Ewe | 36 (11.6) |  | 3 (15.8) | 33 (11.3) |  |  | 23 (11.4) | 10 (11.1) |  |
| Fantse | 21 (6.8) |  | 10 (52.6) | 137 (47.1) |  |  | 12 (6.0) | 7 (7.8) |  |
| Others | 33 (10.7) |  | 1 (5.3) | 32 (11.0) |  |  | 19 (9.4) | 13 (14.4) |  |
| **Mother’s educational level** | |  |  |  |  |  |  |  |  |
| No formal education | 40 (12.9) |  | 5 (26.2) | 35 (12.0) | 0.003^*^ |  | 22 (11.0) | 13 (14.4) | 0.196 |
| Primary | 49 (15.8) |  | 7 (36.9) | 42 (14.4) |  |  | 25 (12.4) | 17 (18.9) |  |
| Secondary/Higher | 221 (71.3) |  | 7 (36.9) | 214 (73.6) |  |  | 154 (76.6) | 60 (66.7) |  |
| **Partner's educational level** | |  |  |  |  |  |  |  |  |
| No formal education | 43 (13.9) |  | 6 (31.6) | 37 (12.7) | 0.043^*^ |  | 22 (11.0) | 15 (16.7) | 0.374 |
| Primary | 20 (6.5) |  | 2 (10.5) | 18 (6.2) |  |  | 12 (6.0) | 6 (6.7) |  |
| Secondary/Higher | 247 (79.7) |  | 11 (57.9) | 236 (81.1) |  |  | 167 (83.0) | 69 (76.6) |  |
| **Household size** (mean:6.2; sd:2.5) | |  |  |  |  |  |  |  |  |
| 1-5 people | 133 (42.9) |  | 7 (36.8) | 126 (43.3) | 0.348 |  | 93 (46.3) | 33 (36.7) | 0.030^*^ |
| 6-10 people | 164 (52.9) |  | 10 (52.6) | 154 (52.9) |  |  | 104 (51.7) | 50 (55.5) |  |
| 11+ people | 13 (4.2) |  | 2 (10.6) | 11 (3.8) |  |  | 4 (2.0) | 7 (7.8) |  |
| **Parity** (mean:2.3; sd:1.4) |  |  |  |  |  |  |  |  |  |
| 1 | 126 (40.6) |  | 9 (47.4) | 117 (40.2) | 0.102 |  | 78 (38.8) | 39 (43.3) | 0.289 |
| 2-4 | 143 (46.1) |  | 5 (26.3) | 138 (47.4) |  |  | 101 (50.2) | 37 (41.1) |  |
| 5+ | 41 (13.3) |  | 5 (26.3) | 36 (12.4) |  |  | 22 (11.0) | 14 (15.6) |  |
| **Age of child** (mean:6.7; sd:3.5) | |  |  |  |  |  |  |  |  |
| ≤4 months | 101 (32.6) |  | 7 (36.8) | 94 (32.3) | 0.905 |  | 66 (32.8) | 28 (31.1) | 0.255 |
| 5-8 months | 99 (31.9) |  | 6 (31.6) | 93 (32.0) |  |  | 69 (34.4) | 24 (26.7) |  |
| 9-12 months | 110 (35.3) |  | 6 (31.6) | 104 (35.7) |  |  | 66 (32.8) | 38 (42.2) |  |
| **Mother’s occupation** |  |  |  |  |  |  |  |  |  |
| Unemployed | 60 (19.4) |  | 3 (15.8) | 57 (19.6) | 0.644 |  | 36 (17.9) | 21 (23.2) | 0.217 |
| Farmer | 122 (39.4) |  | 11 (57.8) | 111 (38.1) |  |  | 75 (37.3) | 36 (40.0) |  |
| Trader | 66 (21.3) |  | 3 (15.8) | 63 (21.5) |  |  | 48 (23.8) | 15 (16.7) |  |
| Seamstress | 20 (6.5) |  | 0 (0.0) | 20 (6.9) |  |  | 13 (6.5) | 7 (7.8) |  |
| Hairdresser | 17 (5.5) |  | 1 (5.3) | 16 (5.5) |  |  | 10 (5.0) | 6 (6.7) |  |
| Public/civil servant | 11 (3.5) |  | 0 (0.0) | 11 (3.9) |  |  | 11 (5.5) | 0 (0.0) |  |
| Others | 14 (4.5) |  | 1 (5.3) | 13 (4.5) |  |  | 8 (4.0) | 5 (5.6) |  |
| **Partner's main occupation** | |  |  |  |  |  |  |  |  |
| Unemployed | 1 (0.3) |  | 0 (0.00) | 1 (0.3) | 0.298 |  | 1 (0.5) | 0 (0.0) | 0.933 |
| Farmer | 149 (48.1) |  | 13 (68.4) | 136 (46.7) |  |  | 93 (46.27) | 43 (47.7) |  |
| Artisan | 57 (18.4) |  | 4 (21.1) | 53 (18.2) |  |  | 38 (18.91) | 15 (16.6) |  |
| Driver | 43 (13.9) |  | 0 (0.0) | 43 (14.8) |  |  | 29 (14.43) | 14 (15.6) |  |
| Trader | 24 (7.7) |  | 2 (10.5) | 22 (7.6) |  |  | 14 (6.97) | 8 (8.9) |  |
| Public/civil servant | 22 (7.1) |  | 0 (0.0) | 22 (7.6) |  |  | 17 (8.46) | 5 (5.6) |  |
| Others | 14 (4.5) |  | 0 (0.0) | 14 (4.8) |  |  | 9 (4.48) | 5 (5.6) |  |

**^*^** *p-value < 0.05*	***^a^*** *Chi-square/Fisher’s exact test*

**Table 3: Socioeconomic and antenatal characteristics of respondents**

|  |  |  | **Bivariate analysis** | | | | | | |
| --- | --- | --- | --- | --- | --- | --- | --- | --- | --- |
| **Variables** | **Total (%)** |  | **ANC attendance (%)** | | **p-value^a^** |  | **ANC Initiation (%)** | | **p-value^a^** |
|  |  |  | **No**  **(n=19)** | **Yes**  **(n=291)** |  |  | **Early (n=201)** | **Late (n=90)** |  |
| **Mother's average monthly income** | |  |  |  |  |  |  |  |  |
| No income | 60 (19.4) |  | 3 (15.8) | 57 (19.5) | 0.517 |  | 36 (17.9) | 21 (23.4) | 0.017^*^ |
| <GHc 100.0 (<$9.0) | 21(6.8) |  | 1 (5.3) | 20 (6.9) |  |  | 10 (5.0) | 10 (11.1) |  |
| GHc 100-299 ($9-33) | 40 (12.9) |  | 2 (10.5) | 38 (13.1) |  |  | 28 (13.9) | 10 (11.1) |  |
| GHc 300-499 ($34-55) | 35 (11.3) |  | 4 (21.0) | 31 (10.7) |  |  | 23 (11.5) | 8 (8.9) |  |
| GHc 500-1000($56-111) | 30 (9.7) |  | 0 (0.0) | 30 (10.3) |  |  | 28 (13.9) | 2 (2.2) |  |
| Don’t know | 124 (40.0) |  | 9 (47.4) | 115 (39.5) |  |  | 76 (37.8) | 39 (43.3) |  |
| **Mother’s health insurance status** | |  |  |  |  |  |  |  |  |
| No | 26 (8.4) |  | 4 (21.1) | 22 (7.6) | 0.040^*^ |  | 13 (6.5) | 9 (10.0) | 0.292 |
| Yes | 284 (91.6) |  | 15 (78.9) | 269 (92.4) |  |  | 188 (93.5) | 81 (90.0) |  |
| **Household wealth index** |  |  |  |  |  |  |  |  |  |
| Poor | 131 (42.3) |  | 15 (78.9) | 116 (39.8) | 0.003^*^ |  | 75 (37.31) | 41 (45.6) | 0.029^*^ |
| Middle | 78 (25.2) |  | 3 (15.8) | 75 (25.8) |  |  | 47 (23.38) | 28 (31.1) |  |
| Rich | 101 (32.5) |  | 1 (5.3) | 100 (34.4) |  |  | 79 (39.3) | 21 (23.3) |  |
| **Knowledge about ANC and pregnancy** | |  |  |  |  |  |  |  |  |
| Inadequate | 11 (3.5) |  | 10 (52.6) | 1 (0.3) | <0.001^*^ |  | 1 (0.5) | 0 (0.0) | 0.503 |
| Adequate | 299 (96.5) |  | 9 (47.4) | 290 (99.7) |  |  | 200 (99.5) | 90 (100) |  |
| **Place of ANC** |  |  |  |  |  |  |  |  |  |
| CHPS | 209 (71.8) |  | – | 209 (71.8) | – |  | 148 (73.6) | 61 (67.8) | 0.409 |
| Clinic | 5 (1.7) |  | – | 5 (1.7) |  |  | 2 (1) | 3 (3.3) |  |
| Health centre | 69 (23.7) |  | – | 69 (23.7) |  |  | 45 (22.4) | 24 (26.7) |  |
| Hospital/ Polyclinic | 8 (2.8) |  | – | 8 (2.8) |  |  | 6 (3.0) | 2 (2.2) |  |
| **Frequency of ANC visit** |  |  |  |  |  |  |  |  |  |
| <4 visits | 20 (6.9) |  | 19 (100.0) | 20 (6.9) | – |  | 6 (3.0) | 14 (15.6) | <0.001^*^ |
| ≥4 visits | 271 (93.1) |  |  | 271 (93.1) |  |  | 195 (97.0) | 76 (84.4) |  |
| **Accompanied for ANC visit (at least one)** | | |  |  |  |  |  |  |  |
| No one (Alone) | 198 (68.0) |  | – | 198 (68.0) | – |  | 139 (69.1) | 59 (65.6) | 0.543 |
| Husband/partner/ other | 93 (32.0) |  | – | 93 (32.0) |  |  | 62 (30.9) | 31 (34.4) |  |
| **ANC service provider** |  |  |  |  |  |  |  |  |  |
| CHN/CHO | 7 (2.4) |  | – | 7 (2.4) | – |  | 6 (3.0) | 1 (1.1) | 0.335 |
| Midwife | 284 (97.6) |  | – | 284 (97.6) |  |  | 195 (97.0) | 89 (98.9) |  |
| **Pay for ANC service** |  |  |  |  |  |  |  |  |  |
| No | 193 (66.3) |  | – | 193 (66.3) | – |  | 131 (65.2) | 62 (68.9) | 0.535 |
| Yes | 98 (33.7) |  | – | 98 (33.7) |  |  | 70 (34.8) | 28 (31.1) |  |
| **Average amount spent per ANC visit** | | |  |  |  |  |  |  |  |
| GHc 1.0-5.0 ($0.1-0.5) | 26 (26.5) |  | – | 26 (26.5) | – |  | 17 (24.29) | 9 (32.1) | 0.445 |
| GHc 6.0-10.0 ($0.6-1.1) | 24 (24.5) |  | – | 24 (24.5) |  |  | 17 (24.29) | 7 (25) |  |
| GHc 11.0-15.0 ($1.2-1.7) | 11 (11.2) |  | – | 11 (11.2) |  |  | 6 (8.57) | 5 (17.9) |  |
| GHc 16.0-20.0 ($1.8-2.2) | 20 (20.4) |  | – | 20 (20.4) |  |  | 16 (22.86) | 4 (14.3) |  |
| >GHc 20.0 (>$2.2) | 17 (17.4) |  | – | 17 (17.4) |  |  | 14 (20) | 3 (10.7) |  |
| **Decision maker before attending ANC** | |  |  |  |  |  |  |  |  |
| Self | 240 (77.4) |  | 17 (89.5) | 223 (76.6) | 0.195 |  | 151 (75.1) | 72 (80.0) | 0.364 |
| Husband/partner | 70 (12.6) |  | 2 (10.5) | 68 (23.4) |  |  | 50 (24.9) | 18 (20.0) |  |
| **Place of delivery** |  |  |  |  |  |  |  |  |  |
| Home | 61 (19.7) |  | 15 (78.9) | 46 (15.8) | <0.001^*^ |  | 25 (12.4) | 21 (23.3) | 0.119 |
| Health facility | 249 (80.3) |  | 4 (21.1) | 245 (84.2) |  |  | 176 (87.6) | 69 (76.7) |  |
| **Received home visit from CHPS zone staff** | | |  |  |  |  |  |  |  |
| No | 202 (65.2) |  | 7 (36.8) | 195 (67.0) | 0.007^*^ |  | 142 (70.6) | 53 (58.9) | 0.039^*^ |
| Yes | 108 (34.8) |  | 12 (63.2) | 96 (33.0) |  |  | 59 (29.4) | 37 (41.1) |  |

**^*^** *p-value < 0.05*	***^a^*** *Chi-square/Fisher’s exact test*

**Table 4: Distance and location of CHPS zones characteristics**

|  |  |  | **Bivariate analysis** | | | | | | |
| --- | --- | --- | --- | --- | --- | --- | --- | --- | --- |
| **Variables** | **Total (%)** |  | **ANC attendance (%)** | | **p-value^a^** |  | **ANC Initiation (%)** | | **p-value^a^** |
|  |  |  | **No**  **(n=19)** | **Yes**  **(n=291)** |  |  | **Early (n=201)** | **Late (n=90)** |  |
| **Health facility in community** | |  |  |  |  |  |  |  |  |
| No | 163 (52.6) |  | 18 (94.7) | 145 (49.8) | <0.001^*^ |  | 98 (48.8) | 47 (52.2) | 0.585 |
| Yes | 147 (47.4) |  | 1 (5.3) | 146 (50.2) |  |  | 103 (51.2) | 43 (47.8) |  |
| **Type of nearest health facility in community** | |  |  |  |  |  |  |  |  |
| CHPS | 250 (80.7) |  | 18 (94.7) | 232 (79.7) | 0.268 |  | 155 (77.1) | 77 (85.6) | 0.251 |
| Health centre | 10 (3.2) |  | 1 (5.3) | 49 (16.8) |  |  | 38 (18.9) | 11 (12.2) |  |
| Clinic | 50 (16.1) |  | 0 (0.00) | 10 (3.5) |  |  | 8 (4.0) | 2 (2.2) |  |
| **CHPS zone functionality status** | |  |  |  |  |  |  |  |  |
| Uncompleted functional | 48 (15.5) |  | 7 (36.8) | 41 (14.1) | 0.008^*^ |  | 20 (10.0) | 21 (23.3) | 0.002^*^ |
| Completed functional | 262 (84.5) |  | 12 (63.2) | 250 (85.9) |  |  | 181 (90.0) | 69 (76.7) |  |
| **Average travel time to nearest CHPS zone (one-way)** | | | |  |  |  |  |  |  |
| ≤30 min | 253 (81.6) |  | 13 (68.4) | 240 (82.5) | 0.126 |  | 171 (85.1) | 69 (76.7) | 0.081 |
| >30 min | 57 (18.4) |  | 6 (31.6) | 51 (17.5) |  |  | 30 (14.9) | 21 (23.3) |  |
| **Means of transport to nearest CHPS zone** | | |  |  |  |  |  |  |  |
| Taxi/bus | 58 (18.7) |  | 16 (84.2) | 264 (90.7) | 0.593 |  | 185 (92.0) | 79 (87.8) | 0.342 |
| Motorbike/tricycle | 108 (34.8) |  | 3 (15.8) | 26 (8.9) |  |  | 15 (7.5) | 11 (12.2) |  |
| Walk | 144 (46.5) |  | 0 (0.0) | 1 (0.34) |  |  | 1 (0.5) | 0 (0.0) |  |
| **Average transportation cost per visit** | |  |  |  |  |  |  |  |  |
| GHc 1-10.0 ($0.1-1.1) | 280 (90.3) |  | 16 (84.2) | 264 (90.7) | 0.593 |  | 185 (92.0) | 79 (87.8) | 0.342 |
| GHc 11-20.0 ($1.2-2.2) | 29 (9.4) |  | 3 (15.8) | 26 (8.9) |  |  | 15 (7.5) | 11 (12.2) |  |
| >GHc 20.0 (>$2.2) | 1 (0.3) |  | 0 (0.0) | 1 (0.4) |  |  | 1 (0.50) | 0 (0.0) |  |
| **Condition of road to health facility** | |  |  |  |  |  |  |  |  |
| No road/Footpath | 9 (2.9) |  | 2 (10.5) | 7 (2.4) | 0.212 |  | 6 (3.0) | 1 (1.1) | 0.233 |
| Feeder/dusty road | 174 (56.1) |  | 11 (57.9) | 163 (56.0) |  |  | 110 (54.7) | 53 (58.9) |  |
| Pothole riddled road | 23 (7.4) |  | 1 (5.3) | 22 (7.6) |  |  | 12 (6.0) | 10 (11.1) |  |
| Smooth tarred road | 104 (33.6) |  | 5 (26.3) | 99 (34.0) |  |  | 73 (36.3) | 26 (28.9) |  |
| **Distance to nearest functional CHPS zones** (mean:2.0; sd:1.5) | | | | |  |  |  |  |  |
| ≥5km | 60 (19.3) |  | 7 (36.8) | 53 (18.2) | 0.046^*^ |  | 29 (14.4) | 24 (26.7) | 0.012^*^ |
| <5km | 250 (80.7) |  | 12 (63.2) | 238 (81.8) |  |  | 172 (85.6) | 66 (73.3) |  |

**^*^***p-value < 0.05*	***^a^*** *Chi-square/Fisher’s exact test*

**Table 5: Logistic regression on associated factors for antenatal accessibility**

| **Logistic regression model** | **COR (95% CI)** | **p-value** |  | **AOR (95% CI)** | **p-value** |
| --- | --- | --- | --- | --- | --- |
| ***Logistic regression for ‘At least one ANC attendance’*** | |  |  |  |  |
| **Marital status** |  |  |  |  |  |
| Not married | 0.357 (0.138, 0.927) | 0.034^*^ |  | **0.125 (0.012, 0.926)** | **0.047^*^** |
| Married | Reference | | | | |
| **Mother’s educational level** |  |  |  |  |  |
| No formal education | 0.229 (0.069, 0.762) | 0.016^*^ |  | 2.474 (0.081, 75.18) | 0.603 |
| Primary | 0.196 (0.065, 0.589) | 0.004^*^ |  | 2.325 (0.134, 40.48) | 0.563 |
| Secondary/Higher | Reference | | | | |
| **Partner's educational level** |  |  |  |  |  |
| No formal education | 0.287 (0.100, 0.824) | 0.020^*^ |  | 0.445 (0.158, 12.56) | 0.635 |
| Primary | 0.419 (0.086, 2.039) | 0.282 |  | 0.922 (0.051, 16.60) | 0.956 |
| Secondary/Higher | Reference | | | | |
| **Mother’s health insurance status** |  |  |  |  |  |
| No | 0.307 (0.093, 0.993) | 0.041^*^ |  | 0.962 (0.068, 13.54) | 0.977 |
| Yes | Reference | | | | |
| **Household wealth index** |  |  |  |  |  |
| Poor | 0.077 (0.010, 0.596) | 0.014^*^ |  | 0.001 (0.0003, 1.250) | 0.934 |
| Middle | 0.250 (0.025, 2.451) | 0.234 |  | 0.009 (0.001, 2.201) | 0.992 |
| Rich | Reference | | | | |
| **Knowledge about ANC and pregnancy** |  |  |  |  |  |
| Inadequate | 0.003 (0.0004, 0.027) | <0.001^*^ |  | 0.006 (0.0001, 1.152) | 0.989 |
| Adequate | Reference | | | | |
| **Place of delivery** |  |  |  |  |  |
| Home | 0.050 (0.016, 0.158) | <0.001^*^ |  | **0.013 (0.001, 0.176)** | **0.001^*^** |
| Health facility | Reference | | | | |
| **Received home visit from CHPS zone staff** |  |  |  |  |  |
| Yes | 0.287 (0.110, 0.753) | 0.011^*^ |  | 0.873 (0.281, 12.49) | 0.517 |
| No | Reference | | | | |
| **Health facility in community** |  |  |  |  |  |
| No | 0.055 (0.007, 0.419) | 0.005^*^ |  | 0.171 (0.013, 2.302) | 0.183 |
| Yes | Reference | | | | |
| **CHPS zone functionality status** |  |  |  |  |  |
| Uncompleted functional | 0.281 (0.105, 0.756) | 0.012^*^ |  | 0.203 (0.017, 2.409) | 0.206 |
| Completed functional | Reference | | | | |
| **Distance to nearest functional CHPS zones** |  |  |  |  |  |
| ≥5km | 0.382 (0.143, 0.902) | 0.044^*^ |  | 0.664 (0.068, 6.441) | 0.724 |
| <5km | Reference | | | | |
| ***Regression model*** | *R^2^ = 0.690 p-value <0.001* | | | | |
| ***Logistic regression for ‘Late ANC initiation’*** | |  |  |  |  |
| **Family size** |  |  |  |  |  |
| 1-5 people | 0.738 (0.438, 1.243) | 0.253 |  | 0.956 (0.537, 1.703) | 0.878 |
| 6-10 people | Reference | | | | |
| 11+ people | 3.640 (1.018, 13.01) | 0.047^*^ |  | **3.848 (1.914, 16.21)** | **0.046^*^** |
| **Mother's average monthly income** |  |  |  |  |  |
| No income | Reference | | | | |
| <GHc 100.0 (<$9.0) | 1.714 (0.613, 4.794) | 0.304 |  | 1. 227 (0.387, 3.889) | 0.728 |
| GHc 100-299 ($9-33) | 0.612 (0.249, 1.507) | 0.286 |  | 0.641 (0.242, 1.697) | 0.371 |
| GHc 300-499 ($34-55) | 0.974 (0.330, 2.871) | 0.295 |  | 0.617 (0.222, 1.713) | 0.354 |
| GHc 500-1000 ($56-111) | 0.122 (0.026, 0.567) | 0.007^*^ |  | **0.123 (0.024, 0.630)** | **0.012^*^** |
| Don’t know | 0.880 (0.454, 1.706) | 0.704 |  | 0.795 (0.387, 1.633) | 0.533 |
| **Household wealth index** |  |  |  |  |  |
| Poor | 2.057 (1.113, 3.798) | 0.021^*^ |  | 1.993 (0.989, 3.947) | 0.058 |
| Middle | 2.241 (1.145, 4.384) | 0.018^*^ |  | 2.032 (0.971, 4.248) | 0.060 |
| Rich | Reference | | | | |
| **Frequency of ANC visit** |  |  |  |  |  |
| <4 visits | 6.987 (2.219, 16.15) | <0.001^*^ |  | **6.332 (2.049, 19.57)** | **0.001^*^** |
| ≥4 visits | Reference | | | | |
| **Received home visit from CHPS zone staff** |  |  |  |  |  |
| Yes | 1.680 (1.001, 2.821) | 0.040^*^ |  | **1.813 (1.014, 3.243)** | **0.045^*^** |
| No | Reference | | | | |
| **CHPS zone functionality status** |  |  |  |  |  |
| Uncompleted functional | 2.754 (1.406, 5.395) | 0.003^*^ |  | 1.647 (0.693, 3.912) | 0.259 |
| Completed functional | Reference | | | | |
| **Distance to nearest functional CHPS zones** |  |  |  |  |  |
| ≥5km | 2.157 (1.171, 3.972) | 0.014^*^ |  | 1.520 (0.691, 3.341) | 0.297 |
| <5km | Reference | | | | |
| ***Regression model*** | *R^2^ = 0.490 p-value <0.001* | | | | |

**^*^***p-value < 0.05*
